# Neutrophil-to-Lymphocyte Ratio Identifies a Pulmonary Vascular High-Risk Phenotype in Heart Failure with Preserved Ejection Fraction

**DOI:** 10.64898/2026.09.08.26362576

**Authors:** Maor Bril, Elchanan Parnasa, Akram Sufian, Khalid Kamal, Fadel Bahouth, Rabea Asleh

## Abstract

**Background:** Pulmonary vascular disease identifies an important phenotype of heart failure with preserved ejection fraction (HFpEF), but the relationship between systemic leukocyte activation and invasively measured pulmonary vascular hemodynamics is incompletely understood. We investigated whether the neutrophil-to-lymphocyte ratio (NLR) is associated with combined post- and precapillary pulmonary hypertension (CpcPH), pulmonary vascular burden, and subsequent cardiovascular events in HFpEF.

**Methods:** We retrospectively studied 169 adults with HFpEF and invasively confirmed postcapillary pulmonary hypertension undergoing clinically indicated right heart catheterization. CpcPH was defined as pulmonary vascular resistance (PVR) >2 Wood units. Associations between NLR and CpcPH were evaluated using multivariable logistic regression. Fine-Gray competing-risk models assessed the composite of cardiovascular death or first heart failure hospitalization.

**Results:** Overall, 80 patients had isolated postcapillary pulmonary hypertension and 89 had CpcPH. PVR increased across ascending NLR tertiles (1.8 [1.4-2.5], 2.2 [1.6-3.1], and 2.6 [1.7-3.8] Wood units; P for trend=0.002), whereas pulmonary artery wedge pressure showed no significant trend. Compared with the lowest NLR tertile, the highest tertile was associated with greater odds of CpcPH after adjustment for clinical covariates and filling pressure (adjusted odds ratio, 2.47 [95% CI, 1.01-6.01]; P=0.047). Higher NLR tertiles were also associated with a graded increase in cardiovascular events (Gray’s P<0.001). The highest NLR tertile remained associated with outcome after adjustment for clinical and hemodynamic variables (adjusted subdistribution hazard ratio [sHR], 2.37 [95% CI, 1.12-5.03]; P=0.024).

**Conclusions:** Among patients with HFpEF and postcapillary pulmonary hypertension, higher NLR was associated with greater pulmonary vascular hemodynamic burden, CpcPH, and subsequent cardiovascular risk. NLR may provide a readily available marker of a high-risk HFpEF phenotype warranting prospective validation.

**Clinical Perspective:** *What Is New?:*

- In patients with HFpEF and invasively confirmed postcapillary pulmonary hypertension, higher neutrophil-to-lymphocyte ratio (NLR) was associated with progressively greater pulmonary vascular resistance without a corresponding significant increase in left-sided filling pressure.
- Higher NLR was associated with greater odds of combined post- and precapillary pulmonary hypertension and with increased risk of cardiovascular death or first heart failure hospitalization, with the prognostic association persisting after adjustment for clinical and invasive hemodynamic variables.

*What Are the Clinical Implications?:*

- NLR, a readily available blood-count-derived marker, may complement invasive hemodynamic phenotyping by identifying patients with HFpEF-associated pulmonary hypertension who have greater pulmonary vascular burden and higher cardiovascular risk, although prospective validation is required before its use for clinical decision-making.

## Introduction

Heart failure with preserved ejection fraction (HFpEF) is a heterogeneous clinical syndrome in which systemic comorbidities, inflammation, endothelial dysfunction, and abnormalities in cardiovascular reserve converge to produce distinct pathophysiological phenotypes (1–3). Pulmonary hypertension (PH) is among the most consequential manifestations of advanced HFpEF, contributing to exercise intolerance, right ventricular dysfunction, and adverse clinical outcomes (4,5). Although PH initially reflects backward transmission of elevated left-sided filling pressures, a subset of patients develops a superimposed precapillary component characterized by increased pulmonary vascular resistance (PVR) (6). Contemporary hemodynamic definitions distinguish isolated postcapillary PH (IpcPH) from combined post- and precapillary PH (CpcPH), defined by PVR >2 Wood units in the presence of postcapillary PH (6,7). The emergence of CpcPH identifies pulmonary vascular disease that is not fully accounted for by passive venous congestion and represents an important target for improved biological and clinical phenotyping (5,8,9).

Systemic inflammation is increasingly recognized as a major pathobiological component of HFpEF (2,10). Comorbidity-driven inflammatory signaling may promote endothelial dysfunction, impaired nitric oxide (NO) bioavailability, oxidative stress, and vascular dysfunction, providing a plausible biological link between systemic inflammatory burden and pulmonary vascular disease (4,11). However, clinically accessible biomarkers capable of identifying this inflammatory-pulmonary vascular interface remain poorly defined. C-reactive protein (CRP), although widely used as a marker of systemic inflammation, reflects hepatic acute-phase signaling and has shown variable prognostic associations in HFpEF (12,13). In contrast, the neutrophil-to-lymphocyte ratio (NLR) integrates two complementary components of systemic immune and physiological stress-relative neutrophilia reflecting innate immune activation and relative lymphopenia associated with neurohormonal stress and impaired adaptive immune regulation (14). Higher NLR has been associated with mortality and heart failure hospitalization across heart failure populations, including HFpEF (15–17), and preliminary observations suggest an association between NLR and increased PVR in patients with heart failure (18). Whether NLR is associated with the invasively defined pulmonary vascular phenotype of CpcPH in HFpEF, independent of the magnitude of left-sided filling pressure, remains uncertain.

Accordingly, we investigated the relationship between NLR, invasive pulmonary hemodynamics, and subsequent cardiovascular outcomes in patients with HFpEF and postcapillary PH undergoing right heart catheterization. We hypothesized that higher NLR would be associated with greater pulmonary vascular hemodynamic burden and a higher likelihood of CpcPH, rather than simply tracking the severity of left-sided congestion, and that this inflammatory signal would identify patients at increased risk of cardiovascular death or heart failure hospitalization. By integrating a routinely available leukocyte-derived biomarker with invasive hemodynamic phenotyping and longitudinal outcomes, we sought to determine whether NLR identifies a clinically accessible high-risk pulmonary vascular phenotype within HFpEF.

## Methods

### Study Design and Population

This retrospective observational cohort study included consecutive adults aged ≥18 years who underwent clinically indicated right heart catheterization (RHC) at Hadassah Medical Center, Israel, between January 2020 and June 2023. Patients were eligible if they had a left ventricular ejection fraction (LVEF) ≥50% and invasive hemodynamic evidence of postcapillary pulmonary hypertension (PH), defined by mean pulmonary artery pressure (mPAP) >20 mmHg and pulmonary artery wedge pressure (PAWP) >15 mmHg at rest, consistent with contemporary hemodynamic definitions (5,6).

To minimize phenotypic heterogeneity and enrich the cohort for HFpEF-associated PH, predefined exclusion criteria were applied. Patients with cardiac amyloidosis, hypertrophic or restrictive cardiomyopathy, constrictive pericarditis, severe aortic valvular disease, prior heart transplantation, congenital heart disease, severe chronic lung disease, pulmonary arterial hypertension, or chronic thromboembolic PH were excluded. Because the primary exposure was derived from circulating leukocyte counts, patients with hematologic disorders expected to substantially alter leukocyte profiles, including acute myeloid leukemia, myelodysplastic syndromes, myeloproliferative neoplasms, multiple myeloma, and Waldenström macroglobulinemia, were also excluded.

The study was conducted in accordance with the Declaration of Helsinki and was approved by the Hadassah Medical Center Institutional Review Board (HMO-0071-26). Because of the retrospective nature of the study, the requirement for individual informed consent was waived by the IRB. The data underlying this study are not publicly available because of patient privacy and institutional restrictions. Deidentified data may be made available from the corresponding author upon reasonable request and subject to institutional and regulatory approval.

### Clinical and Laboratory Assessment

Demographic characteristics, clinical status, comorbidities, laboratory measurements, and medical therapies were systematically extracted from the institutional electronic medical record. Clinical variables included age, sex, body mass index, New York Heart Association functional class, and the H₂FPEF score (19). Comorbidities included hypertension, diabetes mellitus, ischemic heart disease, atrial fibrillation, chronic kidney disease, obstructive sleep apnea, and systemic inflammatory disease. Cardiovascular therapies included renin-angiotensin-aldosterone system inhibitors, β-blockers, mineralocorticoid receptor antagonists, loop diuretics, and sodium-glucose cotransporter 2 inhibitors.

Laboratory measurements obtained contemporaneously with the index RHC were used for the primary analyses. The NLR was calculated by dividing the absolute neutrophil count by the absolute lymphocyte count obtained from a standard complete blood count with differential. Renal function was estimated using the 2021 CKD-EPI creatinine-based equation (20). CRP concentrations were recorded when clinically available and were used in a prespecified exploratory biomarker analysis.

### Invasive Hemodynamic Assessment

RHC was performed for clinical indications using standard balloon-tipped pulmonary artery catheters. Right atrial pressure (RAP), systolic and diastolic pulmonary artery pressures, mPAP, and PAWP were measured at end-expiration according to standard invasive hemodynamic practice (21,22). Cardiac output was primarily determined using the estimated Fick method; thermodilution-derived cardiac output was used when Fick-derived measurements were unavailable. Cardiac index was calculated by indexing cardiac output to body surface area. The transpulmonary pressure gradient (TPG) was calculated as the difference between mPAP and PAWP, and pulmonary vascular resistance (PVR) as: PVR = (mPAP − PAWP) / cardiac output and expressed in Wood units. Pulmonary artery compliance was calculated as stroke volume divided by pulmonary artery pulse pressure. Pulmonary artery pulsatility index was calculated using standard methodology as the pulmonary artery pulse pressure divided by RAP (21).

### Echocardiographic Assessment

Transthoracic echocardiographic data obtained in temporal proximity to the index RHC were extracted from the clinical record. Variables included LVEF, left atrial dimension, average mitral E/e′ ratio calculated from septal and lateral mitral annular tissue Doppler velocities, and estimated right ventricular systolic pressure.

### Hemodynamic Phenotype

Patients with postcapillary PH were classified according to the 2022 ESC/ERS hemodynamic framework (6). IpcPH was defined by PAWP >15 mmHg and PVR ≤2 Wood units, whereas CpcPH was defined by PAWP >15 mmHg and PVR >2 Wood units. The latter identifies a precapillary hemodynamic component superimposed on elevated left-sided filling pressure. The primary hemodynamic analysis evaluated the association between NLR and CpcPH. Because CpcPH is defined directly by a PVR threshold, PVR was additionally examined as a continuous measure of pulmonary vascular load across the NLR distribution.

### Clinical Outcomes

The primary clinical outcome was the composite of cardiovascular death or first hospitalization for heart failure occurring during follow-up after the index RHC. Heart failure hospitalization was defined as an unplanned hospitalization requiring signs/symptoms of worsening HF and intensification of HF therapy. Clinical outcomes were ascertained from institutional records or the national mortality registry, with follow-up beginning on the date of the index RHC and continuing until the first relevant outcome, death, loss to follow-up, or last follow-up, whichever occurred first.

### Statistical Analysis

Continuous variables were assessed for distribution and are presented as mean ± SD when approximately normally distributed and median (interquartile range) when skewed. Categorical variables are presented as number (percentage). Between-group comparisons were performed using the Student’s t-test or Mann-Whitney U test for continuous variables and the χ² or Fisher exact test for categorical variables, as appropriate. NLR was evaluated both continuously and in cohort-derived tertiles. The continuous analysis preserved information across the full NLR distribution, whereas tertiles were used to facilitate clinical interpretation and to characterize graded differences in hemodynamic phenotype. Potential nonlinear associations between continuous NLR and CpcPH were evaluated using restricted cubic splines with 3 knots. Hemodynamic variables across ascending NLR tertiles were compared using analysis of variance or the Kruskal–Wallis test, as appropriate; ordered trends were assessed using the Jonckheere–Terpstra test.

Associations between NLR and the CpcPH phenotype were evaluated using logistic regression and are reported as odds ratios (ORs) with 95% confidence intervals (CIs). Multivariable covariates were selected a priori based on clinical relevance and their potential relationships with systemic inflammatory status and pulmonary vascular hemodynamics rather than solely on univariable statistical significance. The clinical model included age, sex, body mass index, atrial fibrillation, and estimated glomerular filtration rate. A subsequent model additionally accounted for left-sided filling pressure. Additionally, multivariable linear regression was used to evaluate the association between continuous NLR and PVR, utilizing the same clinical and hemodynamic covariates.

For longitudinal analyses, cumulative incidence functions were used to estimate the incidence of the primary cardiovascular outcome across NLR tertiles, with non-cardiovascular death treated as a competing event; distributions were compared using Gray’s test. Fine–Gray proportional subdistribution hazards regression was used to estimate subdistribution hazard ratios (sHRs) and 95% CIs for cardiovascular death or first heart failure hospitalization. NLR was evaluated both continuously and categorically, with the lowest tertile serving as the reference group. Furthermore, multivariable outcome models were constructed to determine whether NLR provided prognostic information beyond relevant clinical and invasive hemodynamic characteristics. Missing covariate data were addressed using multiple imputation by chained equations with 10 imputed data sets, and regression estimates were pooled according to Rubin’s rules. Multicollinearity was evaluated using variance inflation factors.

Prespecified sensitivity analyses included assessment of effect modification by biological sex, re-evaluation of the CpcPH outcome using a stricter hemodynamic threshold (PVR >3 Wood units), and an exploratory comparison of log-transformed NLR and CRP among patients with available baseline CRP measurements. Spearman rank correlation was used to quantify the relationship between NLR and CRP.

All tests were 2-sided, with P<0.05 considered statistically significant. Analyses were performed using R software version 4.4.1 (R Foundation for Statistical Computing). The authors had full access to the data in the study and take responsibility for the integrity of the data and the accuracy of the data analysis

## Results

### Study Population and Baseline Characteristics

The final study cohort included 169 patients with HFpEF and invasively confirmed postcapillary pulmonary hypertension, of whom 80 met criteria for IpcPH and 89 for CpcPH. The mean age was 71.4±10.5 years, and 62% were women. Baseline clinical and hemodynamic characteristics according to pulmonary hypertension phenotype are presented in ***Table 1***. Compared with patients with IpcPH, those with CpcPH were older (73.9±9.8 versus 68.6±10.6 years; P=0.001), had lower body mass index (30.0±6.3 versus 33.2±7.8 kg/m²; P=0.007), and more frequently had an H₂FPEF score ≥6 (66% versus 40%; P<0.001) and atrial fibrillation (62% versus 38%; P=0.002).

**Table 1.**
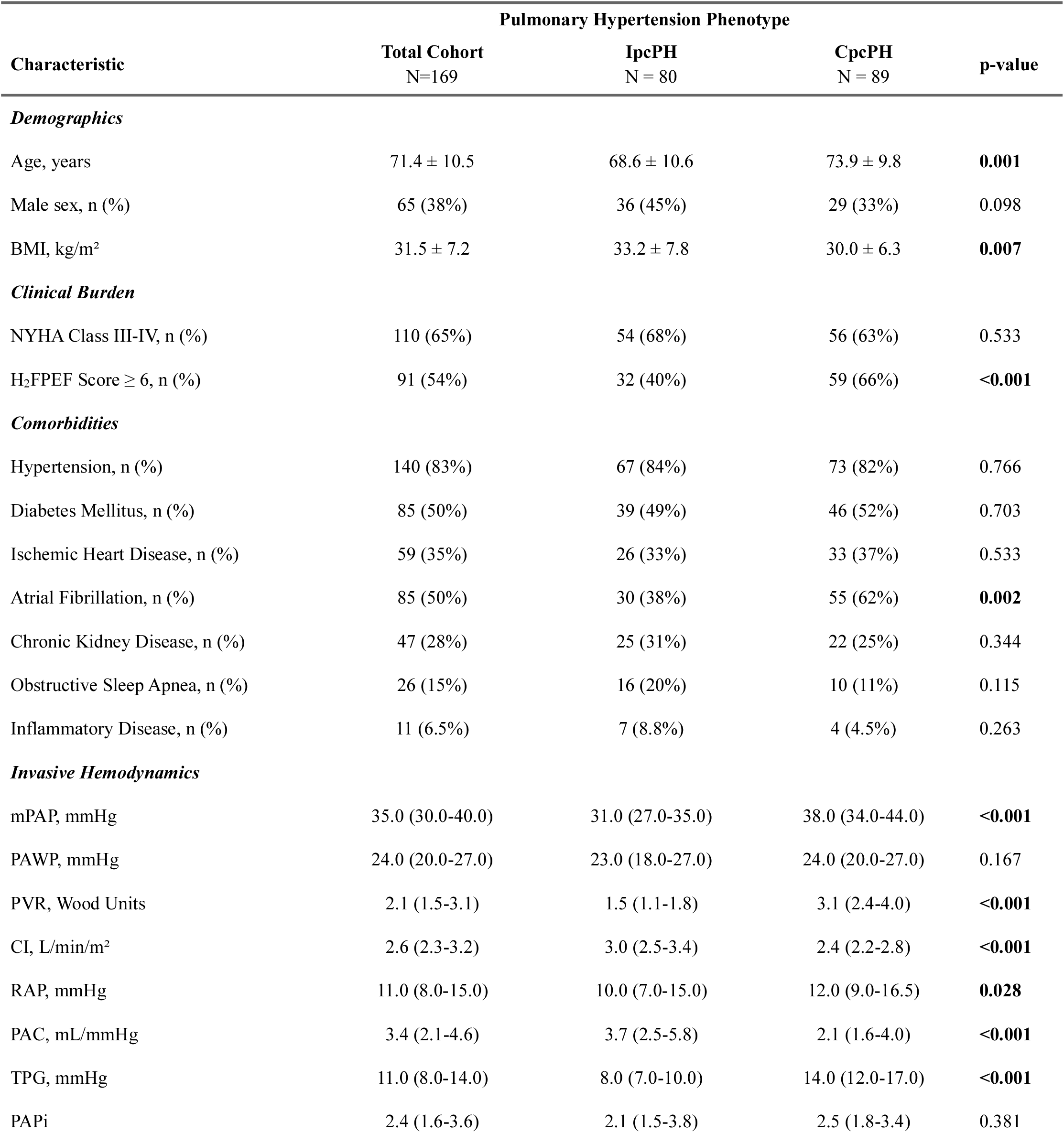
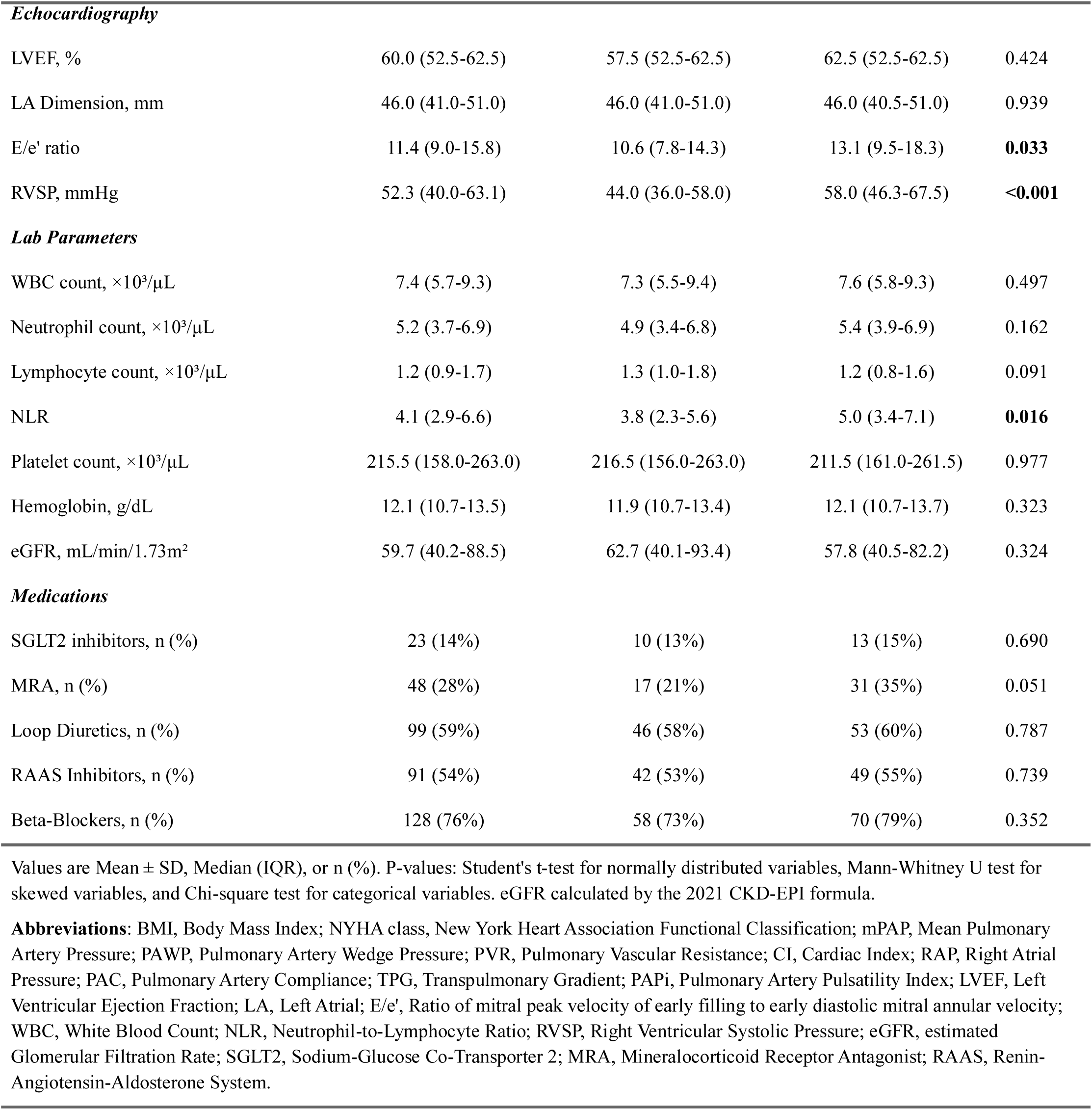
Baseline Characteristics by Pulmonary Hypertension Phenotype.

|  | Pulmonary Hypertension Phenotype |  |  |  |
| --- | --- | --- | --- | --- |
| Characteristic | Total Cohort<br>N=169 | IpcPH<br>N = 80 | CpcPH<br>N = 89 | p-value |
| *Demographics* |  |  |  |  |
| Age, years | 71.4 ± 10.5 | 68.6 ± 10.6 | 73.9 ± 9.8 | **0.001** |
| Male sex, n (%) | 65 (38%) | 36 (45%) | 29 (33%) | 0.098 |
| BMI, kg/m² | 31.5 ± 7.2 | 33.2 ± 7.8 | 30.0 ± 6.3 | **0.007** |
| *Clinical Burden* |  |  |  |  |
| NYHA Class III-IV, n (%) | 110 (65%) | 54 (68%) | 56 (63%) | 0.533 |
| H<sub>2</sub>FPEF Score ≥ 6, n (%) | 91 (54%) | 32 (40%) | 59 (66%) | **<0.001** |
| *Comorbidities* |  |  |  |  |
| Hypertension, n (%) | 140 (83%) | 67 (84%) | 73 (82%) | 0.766 |
| Diabetes Mellitus, n (%) | 85 (50%) | 39 (49%) | 46 (52%) | 0.703 |
| Ischemic Heart Disease, n (%) | 59 (35%) | 26 (33%) | 33 (37%) | 0.533 |
| Atrial Fibrillation, n (%) | 85 (50%) | 30 (38%) | 55 (62%) | **0.002** |
| Chronic Kidney Disease, n (%) | 47 (28%) | 25 (31%) | 22 (25%) | 0.344 |
| Obstructive Sleep Apnea, n (%) | 26 (15%) | 16 (20%) | 10 (11%) | 0.115 |
| Inflammatory Disease, n (%) | 11 (6.5%) | 7 (8.8%) | 4 (4.5%) | 0.263 |
| *Invasive Hemodynamics* |  |  |  |  |
| mPAP, mmHg | 35.0 (30.0-40.0) | 31.0 (27.0-35.0) | 38.0 (34.0-44.0) | **<0.001** |
| PAWP, mmHg | 24.0 (20.0-27.0) | 23.0 (18.0-27.0) | 24.0 (20.0-27.0) | 0.167 |
| PVR, Wood Units | 2.1 (1.5-3.1) | 1.5 (1.1-1.8) | 3.1 (2.4-4.0) | **<0.001** |
| CI, L/min/m² | 2.6 (2.3-3.2) | 3.0 (2.5-3.4) | 2.4 (2.2-2.8) | **<0.001** |
| RAP, mmHg | 11.0 (8.0-15.0) | 10.0 (7.0-15.0) | 12.0 (9.0-16.5) | **0.028** |
| PAC, mL/mmHg | 3.4 (2.1-4.6) | 3.7 (2.5-5.8) | 2.1 (1.6-4.0) | **<0.001** |
| TPG, mmHg | 11.0 (8.0-14.0) | 8.0 (7.0-10.0) | 14.0 (12.0-17.0) | **<0.001** |
| PAPi | 2.4 (1.6-3.6) | 2.1 (1.5-3.8) | 2.5 (1.8-3.4) | 0.381 |
| Characteristic | Total Cohort<br>N=169 | IpcPH<br>N = 80 | CpcPH<br>N = 89 | p-value |
| *Echocardiography* |  |  |  |  |
| LVEF, % | 60.0 (52.5-62.5) | 57.5 (52.5-62.5) | 62.5 (52.5-62.5) | 0.424 |
| LA Dimension, mm | 46.0 (41.0-51.0) | 46.0 (41.0-51.0) | 46.0 (40.5-51.0) | 0.939 |
| E/e' ratio | 11.4 (9.0-15.8) | 10.6 (7.8-14.3) | 13.1 (9.5-18.3) | **0.033** |
| RVSP, mmHg | 52.3 (40.0-63.1) | 44.0 (36.0-58.0) | 58.0 (46.3-67.5) | **<0.001** |
| *Lab Parameters* |  |  |  |  |
| WBC count, ×10<sup>3</sup>/μL | 7.4 (5.7-9.3) | 7.3 (5.5-9.4) | 7.6 (5.8-9.3) | 0.497 |
| Neutrophil count, ×10<sup>3</sup>/μL | 5.2 (3.7-6.9) | 4.9 (3.4-6.8) | 5.4 (3.9-6.9) | 0.162 |
| Lymphocyte count, ×10<sup>3</sup>/μL | 1.2 (0.9-1.7) | 1.3 (1.0-1.8) | 1.2 (0.8-1.6) | 0.091 |
| NLR | 4.1 (2.9-6.6) | 3.8 (2.3-5.6) | 5.0 (3.4-7.1) | **0.016** |
| Platelet count, ×10<sup>3</sup>/μL | 215.5 (158.0-263.0) | 216.5 (156.0-263.0) | 211.5 (161.0-261.5) | 0.977 |
| Hemoglobin, g/dL | 12.1 (10.7-13.5) | 11.9 (10.7-13.4) | 12.1 (10.7-13.7) | 0.323 |
| eGFR, mL/min/1.73m<sup>2</sup> | 59.7 (40.2-88.5) | 62.7 (40.1-93.4) | 57.8 (40.5-82.2) | 0.324 |
| *Medications* |  |  |  |  |
| SGLT2 inhibitors, n (%) | 23 (14%) | 10 (13%) | 13 (15%) | 0.690 |
| MRA, n (%) | 48 (28%) | 17 (21%) | 31 (35%) | 0.051 |
| Loop Diuretics, n (%) | 99 (59%) | 46 (58%) | 53 (60%) | 0.787 |
| RAAS Inhibitors, n (%) | 91 (54%) | 42 (53%) | 49 (55%) | 0.739 |
| Beta-Blockers, n (%) | 128 (76%) | 58 (73%) | 70 (79%) | 0.352 |

Values are Mean ± SD, Median (IQR), or n (%). P-values: Student's t-test for normally distributed variables, Mann-Whitney U test for skewed variables, and Chi-square test for categorical variables. eGFR calculated by the 2021 CKD-EPI formula.
**Abbreviations:** BMI, Body Mass Index; NYHA class, New York Heart Association Functional Classification; mPAP, Mean Pulmonary Artery Pressure; PAWP, Pulmonary Artery Wedge Pressure; PVR, Pulmonary Vascular Resistance; CI, Cardiac Index; RAP, Right Atrial Pressure; PAC, Pulmonary Artery Compliance; TPG, Transpulmonary Gradient; PAPi, Pulmonary Artery Pulsatility Index; LVEF, Left Ventricular Ejection Fraction; LA, Left Atrial; E/e', Ratio of mitral peak velocity of early filling to early diastolic mitral annular velocity; WBC, White Blood Count; NLR, Neutrophil-to-Lymphocyte Ratio; RVSP, Right Ventricular Systolic Pressure; eGFR, estimated Glomerular Filtration Rate; SGLT2, Sodium-Glucose Co-Transporter 2; MRA, Mineralocorticoid Receptor Antagonist; RAAS, Renin-Angiotensin-Aldosterone System.

Left-sided filling pressure was similar between groups, with a median PAWP of 24.0 [20.0–27.0] mmHg in CpcPH and 23.0 [18.0–27.0] mmHg in IpcPH (P=0.167). In contrast, patients with CpcPH demonstrated greater pulmonary vascular and right-sided hemodynamic burden, including higher PVR (3.1 [2.4–4.0] versus 1.5 [1.1–1.8] Wood units; P<0.001), mPAP (38.0 [34.0–44.0] versus 31.0 [27.0–35.0] mmHg; P<0.001), RAP (12.0 [9.0–16.5] versus 10.0 [7.0–15.0] mmHg; P=0.028), and TPG (14.0 [12.0– 17.0] versus 8.0 [7.0–10.0] mmHg; P<0.001), together with lower cardiac index and pulmonary artery compliance. NLR was higher in patients with CpcPH than in those with IpcPH (5.0 [3.4–7.1] versus 3.8 [2.3–5.6]; P=0.016), whereas total leukocyte and absolute neutrophil counts did not differ significantly between groups.

### NLR and Pulmonary Vascular Hemodynamics

NLR was evaluated both continuously and across cohort-derived tertiles. Restricted cubic spline analysis demonstrated an increase in the probability of CpcPH across the lower-to-intermediate NLR range, with attenuation of the association and greater uncertainty at higher NLR values; however, the test for nonlinearity did not reach statistical significance (P for nonlinearity=0.052) (***Figure 1A***). PVR increased progressively across ascending NLR tertiles, from 1.8 [1.4–2.5] Wood units in tertile 1 to 2.2 [1.6–3.1] in tertile 2 and 2.6 [1.7–3.8] in tertile 3 (P for trend=0.002; ***Figure 1B***). Similar graded increases were observed in RAP (10.0 [6.0–14.0], 12.0 [9.0–16.0], and 12.0 [8.5–17.5] mmHg; P for trend=0.007), mPAP (32.0 [28.0–37.0], 36.0 [29.5–43.5], and 35.0 [32.0–41.5] mmHg; P for trend=0.008), and TPG (10.0 [8.0–13.0], 11.0 [8.0– 16.0], and 12.0 [9.0–16.0] mmHg; P for trend=0.029) (***Figure 2A***; ***Supplementary Table S1***). Conversely, PAWP did not increase significantly across NLR tertiles (21.0 [18.0–26.0], 24.0 [20.0–27.0], and 24.0 [21.0–27.0] mmHg; P for trend=0.079). When PVR was plotted against PAWP, patients with higher NLR values were distributed toward higher PVR across the observed range of filling pressures (***Figure 2B***).

**Figure 1.** Association of Neutrophil-to-Lymphocyte Ratio with Pulmonary Vascular Hemodynamics. (**A**) Restricted cubic spline depicting the association between continuous NLR and the predicted probability of CpcPH. The solid line represents the predicted probability and the shaded band the 95% CI. Background shading denotes cohort-derived NLR tertiles. (**B**) Distribution of PVR across ascending NLR tertiles. Boxes represent the interquartile range, horizontal lines the median, and whiskers 1.5 times the interquartile range. The P value represents the Jonckheere–Terpstra test for ordered trend across NLR tertiles. *Abbreviations*: CpcPH, combined post- and precapillary pulmonary hypertension; NLR, neutrophil-to-lymphocyte ratio; PVR, pulmonary vascular resistance.

**Figure 2.** Pulmonary Vascular and Right-Sided Hemodynamics Across Neutrophil-to-Lymphocyte Ratio Tertiles. (**A**) Distribution of RAP, mPAP, and PAWP across ascending NLR tertiles. P values represent Jonckheere–Terpstra tests for ordered trend. (**B**) Relationship between PVR and PAWP according to NLR. Individual observations are color-coded according to continuous NLR values, and fitted regression lines depict the relationship between PVR and PAWP within each NLR tertile. *Abbreviations*: mPAP, mean pulmonary artery pressure; NLR, neutrophil-to-lymphocyte ratio; PAWP, pulmonary artery wedge pressure; PVR, pulmonary vascular resistance; RAP, right atrial pressure.

### Association Between NLR and CpcPH

Univariable predictors of CpcPH are shown in ***Supplementary Table S2***. In unadjusted logistic regression, patients in the highest NLR tertile (NLR ≥5.55) had higher odds of CpcPH compared with those in the lowest tertile (OR, 2.46 [95% CI, 1.15–5.28]; P=0.021). This association persisted after adjustment for age, sex, body mass index, atrial fibrillation, and estimated glomerular filtration rate (adjusted OR, 2.49 [95% CI, 1.03–6.07]; P=0.044). After additional adjustment for left-sided filling pressure, the association remained similar (adjusted OR, 2.47 [95% CI, 1.01–6.01]; P=0.047) (***Table 2***). The intermediate NLR tertile was not significantly associated with CpcPH in the multivariable models. In the fully adjusted model, male sex and higher body mass index were associated with significantly lower odds of CpcPH, whereas advancing age and atrial fibrillation demonstrated borderline associations.

**Table 2:**
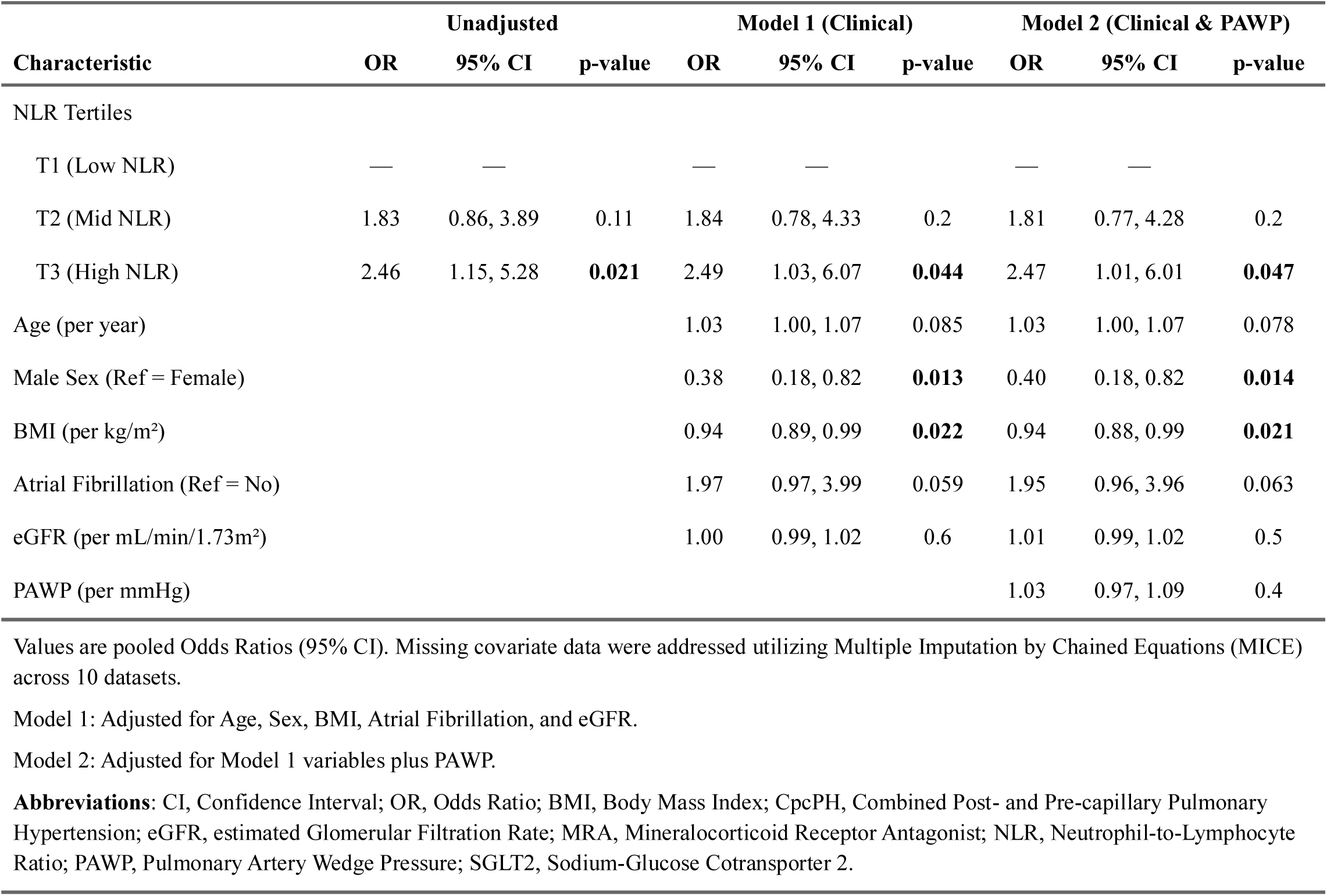
Multivariable Logistic Regression for the Association Between NLR Tertiles and CpcPH.

| Characteristic | Unadjusted |  |  | Model 1 (Clinical) |  |  | Model 2 (Clinical & PAWP) |  |  |
| --- | --- | --- | --- | --- | --- | --- | --- | --- | --- |
|  | OR | 95% CI | p-value | OR | 95% CI | p-value | OR | 95% CI | p-value |
| NLR Tertiles |  |  |  |  |  |  |  |  |  |
| T1 (Low NLR) | — | — |  | — | — |  | — | — |  |
| T2 (Mid NLR) | 1.83 | 0.86, 3.89 | 0.11 | 1.84 | 0.78, 4.33 | 0.2 | 1.81 | 0.77, 4.28 | 0.2 |
| T3 (High NLR) | 2.46 | 1.15, 5.28 | **0.021** | 2.49 | 1.03, 6.07 | **0.044** | 2.47 | 1.01, 6.01 | **0.047** |
| Age (per year) |  |  |  | 1.03 | 1.00, 1.07 | 0.085 | 1.03 | 1.00, 1.07 | 0.078 |
| Male Sex (Ref = Female) |  |  |  | 0.38 | 0.18, 0.82 | **0.013** | 0.40 | 0.18, 0.82 | **0.014** |
| BMI (per kg/m<sup>2</sup>) |  |  |  | 0.94 | 0.89, 0.99 | **0.022** | 0.94 | 0.88, 0.99 | **0.021** |
| Atrial Fibrillation (Ref = No) |  |  |  | 1.97 | 0.97, 3.99 | 0.059 | 1.95 | 0.96, 3.96 | 0.063 |
| eGFR (per mL/min/1.73m<sup>2</sup>) |  |  |  | 1.00 | 0.99, 1.02 | 0.6 | 1.01 | 0.99, 1.02 | 0.5 |
| PAWP (per mmHg) |  |  |  |  |  |  | 1.03 | 0.97, 1.09 | 0.4 |

Values are pooled Odds Ratios (95% CI). Missing covariate data were addressed utilizing Multiple Imputation by Chained Equations (MICE) across 10 datasets.
Model 1: Adjusted for Age, Sex, BMI, Atrial Fibrillation, and eGFR.
Model 2: Adjusted for Model 1 variables plus PAWP.
**Abbreviations:** CI, Confidence Interval; OR, Odds Ratio; BMI, Body Mass Index; CpcPH, Combined Post- and Pre-capillary Pulmonary Hypertension; eGFR, estimated Glomerular Filtration Rate; MRA, Mineralocorticoid Receptor Antagonist; NLR, Neutrophil-to-Lymphocyte Ratio; PAWP, Pulmonary Artery Wedge Pressure; SGLT2, Sodium-Glucose Cotransporter 2.

PVR was additionally evaluated as a continuous outcome. In a multivariable linear regression model, increasing continuous NLR remained a significant independent predictor of higher PVR after adjustment for baseline clinical characteristics and PAWP (β = 0.05 Wood units per 1-unit increase in NLR [95% CI, 0.00–0.10]; P=0.033) (***Supplementary Table S3***). Finally, in a sensitivity analysis applying a stricter historical threshold for CpcPH (PVR >3 Wood units), the highest NLR tertile maintained a strong association with the phenotype (adjusted OR, 6.43 [95% CI, 2.15–21.9]; P=0.002).

### Association Between NLR and Adverse Cardiovascular Outcomes

Over a median follow-up of 46.0 months (95% CI, 41.6–51.0), 64 patients experienced the primary composite outcome of cardiovascular death or first heart failure hospitalization, with cardiovascular death as the first qualifying event in 5 patients and heart failure hospitalization in 59 patients. Eight noncardiovascular deaths occurred during follow-up and were treated as competing events. The cumulative incidence of the primary outcome differed significantly across NLR tertiles (Gray’s P<0.001; ***Figure 3A***). Compared with tertile 1, unadjusted risk was higher in tertile 2 (sHR, 2.31 [95% CI, 1.17–4.58]; P=0.016) and tertile 3 (sHR, 3.66 [95% CI, 1.88–7.15]; P<0.001). NLR was also associated with the primary outcome when modeled continuously (sHR, 1.08 per unit increase [95% CI, 1.05–1.12]; P<0.001) (***Supplementary Table S4***). In contrast, the binary CpcPH phenotype was not significantly associated with the primary composite outcome in unadjusted analysis (sHR, 1.33 [95% CI, 0.81–2.20]; P=0.3).

**Figure 3.** Association of Neutrophil-to-Lymphocyte Ratio with Cardiovascular Outcomes. (**A**) Cumulative incidence functions for the primary composite outcome of cardiovascular death or first heart failure hospitalization during follow-up, stratified by baseline NLR tertiles. Non-cardiovascular death was treated as a competing event. The P value represents Gray’s test for differences among cumulative incidence functions. (**B**) Forest plot showing subdistribution hazard ratios and 95% CIs for the association between NLR and the primary composite outcome derived from multivariable Fine–Gray competing-risk regression models. The model adjusts for age, sex, eGFR, and hypertension, as well as hemodynamic variables, including PVR and RAP. The lowest NLR tertile serves as the reference group. *Abbreviations*: eGFR, estimated glomerular filtration rate; NLR, neutrophil-to-lymphocyte ratio; PAWP, pulmonary artery wedge pressure; PVR, pulmonary vascular resistance; RAP, right atrial pressure.

In the clinical multivariable model, the highest NLR tertile remained significantly associated with the primary outcome after adjustment for baseline clinical characteristics and renal function (adjusted sHR, 3.08 [95% CI, 1.45–6.51]; P=0.003) (***Supplementary Table S5***). This prognostic association was similarly preserved in the hemodynamic model adjusted for PVR and RAP (adjusted sHR, 2.49 [95% CI, 1.23–5.07]; P=0.012). Finally, in the combined clinical and hemodynamic model, the highest NLR tertile remained independently associated with increased cardiovascular risk (adjusted sHR, 2.37 [95% CI, 1.12–5.03]; P=0.024) (***Figure 3B***). Among the other covariates in the combined model, lower eGFR and higher RAP remained independently associated with the primary outcome, whereas PVR did not.

### Sensitivity and Exploratory Analyses

There was no significant interaction between biological sex and the association of NLR tertiles with the primary outcome across the clinical, hemodynamic, or combined multivariable models (all P for interaction >0.05). Among the 110 patients with available baseline CRP measurements, NLR and CRP were weakly correlated (Spearman r=0.27; P=0.005). Baseline CRP concentrations did not differ between patients with IpcPH and CpcPH (P=0.9). In a multivariable Fine–Gray model including both biomarkers, continuous log-transformed NLR remained independently associated with the primary outcome (adjusted sHR, 1.74 per 1 log-unit increase [95% CI, 1.15–2.64]; P=0.008), whereas log-transformed CRP was not associated with cardiovascular events (adjusted sHR, 1.22 per 1 log-unit increase [95% CI, 0.82–1.81]; P=0.3).

## Discussion

In this invasively phenotyped cohort of patients with HFpEF and postcapillary pulmonary hypertension, NLR was associated with pulmonary vascular hemodynamic phenotype and subsequent cardiovascular risk. Several findings are noteworthy. First, higher NLR was associated with progressively greater PVR and a higher likelihood of CpcPH, whereas PAWP did not increase significantly across NLR tertiles. Second, the association between high NLR and CpcPH persisted after adjustment for clinical characteristics and left-sided filling pressure. Third, higher NLR identified a marked gradient in cardiovascular death or heart failure hospitalization and remained associated with adverse outcomes after adjustment for clinical and invasive hemodynamic variables. Finally, the conventional binary classification of CpcPH was not itself significantly associated with outcomes in this cohort, whereas NLR retained a strong prognostic signal. Together, these observations suggest that a routinely available leukocyte-derived index may identify a high-risk HFpEF-PH phenotype characterized by greater pulmonary vascular burden and adverse clinical trajectory.

### NLR and the Pulmonary Vascular Phenotype of HFpEF

Pulmonary hypertension in HFpEF is pathophysiologically heterogeneous. In IpcPH, elevated pulmonary artery pressure predominantly reflects backward transmission of increased left atrial and pulmonary venous pressure. In contrast, CpcPH incorporates an additional precapillary component, conventionally identified by elevated PVR, which may arise from pulmonary vasoconstriction and structural changes within the pulmonary circulation (4,6,8,9). Contemporary frameworks therefore increasingly regard PVR not simply as a classification variable but as an integrated measure of pulmonary vascular load that may identify a biologically distinct stage of HFpEF-PH (5,6,9,21). The recent HFA/ESC scientific statement on PH associated with left heart failure emphasized improved multidimensional phenotyping and rigorous invasive hemodynamic characterization as major priorities for understanding disease heterogeneity and developing phenotype-directed interventions (9).

Against this background, the relationship between NLR and invasive pulmonary vascular hemodynamics represents the principal novel observation of the present study.

Previous studies have established NLR as a marker of adverse prognosis across heart failure populations, including HFpEF (14–17), and preliminary observations have suggested an association between NLR and PVR in unselected patients with congestive heart failure (18). Our findings extend these observations by demonstrating a graded relationship between NLR and pulmonary vascular burden specifically in patients with invasively characterized HFpEF-PH. PVR increased progressively across NLR tertiles, accompanied by increases in TPG, mPAP, and RAP, whereas PAWP showed no statistically significant corresponding trend. Thus, the association between NLR and the pulmonary circulation did not simply parallel increasing left-sided filling pressure.

This distinction is clinically relevant. Invasive physiological studies of PH associated with left heart disease have demonstrated that patients with a precapillary pulmonary vascular component may have similar PAWP to those with IpcPH yet display substantially greater RV-pulmonary arterial uncoupling, impaired cardiac output reserve, and exercise limitation (23). These observations reinforce the concept that pulmonary vascular disease in HFpEF cannot be understood solely from the magnitude of left atrial pressure. Our data suggest that NLR may provide complementary information regarding this pulmonary vascular phenotype. Importantly, however, the present study does not establish that systemic inflammation causes pulmonary vascular remodeling. PVR is a hemodynamic measure and does not directly quantify pulmonary vascular structure. The findings should therefore be interpreted as an association between leukocyte-derived inflammatory burden and pulmonary vascular physiology rather than evidence of a causal inflammatory vasculopathy.

### Biological Links Between Systemic Inflammation and Pulmonary Vascular Dysfunction

The observed association is nevertheless biologically plausible within contemporary models of HFpEF. HFpEF is increasingly viewed as a systemic syndrome in which aging, obesity, metabolic dysfunction, renal disease, and other comorbidities promote chronic inflammatory and endothelial stress (1–3,10,24). These processes have traditionally been examined in relation to coronary microvascular dysfunction, impaired NO–cyclic guanosine monophosphate–protein kinase G signaling, myocardial fibrosis, and increased ventricular stiffness (2,3,11). The pulmonary vascular endothelium is exposed to the same systemic inflammatory environment, while also experiencing chronic mechanical stress from elevated pulmonary venous pressure. The convergence of inflammatory signaling and sustained hemodynamic stress provides a plausible substrate through which pulmonary vascular dysfunction may evolve in susceptible patients (5,8,9).

NLR may be particularly informative because it integrates 2 components of the systemic response rather than representing a single inflammatory mediator. Neutrophilia reflects activation of innate immune pathways and can accompany oxidative stress, endothelial activation, myeloperoxidase release, proteolytic activity, and neutrophil extracellular trap formation, processes capable of altering vascular homeostasis (14,25). Conversely, relative lymphopenia may reflect heightened sympathetic and neurohormonal activation, physiological stress, and impaired adaptive immune regulation (14,16,17). NLR therefore should not be considered a specific marker of neutrophil-mediated pulmonary vascular injury; rather, it may represent an integrated marker of inflammatory, neurohormonal, and systemic stress that tracks with a more advanced cardiovascular phenotype.

This interpretation is also supported by the observation that absolute leukocyte and neutrophil counts did not differ significantly between IpcPH and CpcPH, whereas NLR did. The ratio may therefore capture changes in the balance between innate and adaptive immune compartments that are not apparent from total leukocyte counts alone. At the same time, this finding argues against interpreting NLR as a direct mechanistic biomarker. Whether the association reflects inflammatory activation contributing to pulmonary vascular dysfunction, the systemic consequences of advanced right-sided disease, or bidirectional interactions between congestion and immunity cannot be resolved by the present observational design.

### NLR and Cardiovascular Risk Beyond Hemodynamic Classification

The prognostic findings provide a second clinically relevant dimension to the study. Increasing NLR was associated with a pronounced gradient in cardiovascular death or first heart failure hospitalization, with patients in the highest tertile experiencing the greatest event burden. Importantly, the association persisted after adjustment for clinical markers of disease severity and important hemodynamic parameters, including PVR and RAP. These findings are consistent with previous studies demonstrating the prognostic significance of NLR in HFpEF and broader heart failure populations (14–17,26), but extend them by showing that the prognostic information carried by NLR is not fully captured by invasive pulmonary vascular and right-sided hemodynamics.

Of particular interest, CpcPH as a binary phenotype was not significantly associated with the primary outcome in this cohort, despite the prognostic importance of pulmonary vascular disease established in larger studies (5,6,8). This apparent discrepancy should be interpreted cautiously and may partly reflect limited statistical power and the loss of information inherent in dichotomizing a continuous physiological variable at a single PVR threshold. Nevertheless, it raises an important conceptual point. CpcPH describes the hemodynamic state at the time of RHC, whereas NLR may integrate multiple biological processes, including systemic inflammation, physiological stress, neurohormonal activation, renal dysfunction, and disease acuity, that influence subsequent clinical trajectory. Thus, hemodynamic classification and circulating inflammatory indices may provide complementary rather than interchangeable information.

This concept is particularly relevant to contemporary efforts to move beyond single-variable classification of HFpEF toward multidimensional phenotyping (1,9,27). A patient with a PVR of 2.1 Wood units and one with substantially more severe pulmonary vascular disease are both classified as CpcPH despite potentially very different physiology and clinical risk. Conversely, a circulating biomarker such as NLR cannot define pulmonary vascular disease but may help identify biological heterogeneity within a given hemodynamic category. Integration of readily available biomarkers with quantitative hemodynamics may therefore ultimately prove more informative than either approach alone, although this hypothesis requires validation in larger prospective cohorts.

### NLR Compared With CRP

The exploratory comparison with CRP provides additional context but should be interpreted cautiously. Among patients with available CRP measurements, the correlation between NLR and CRP was weak, suggesting that the 2 measures capture only partially overlapping biological information. CRP was not associated with either CpcPH or cardiovascular events, whereas NLR demonstrated a significant prognostic association in the joint biomarker model.

These findings should not be interpreted as evidence that NLR is biologically more specific than CRP. CRP primarily reflects hepatic acute-phase signaling driven by circulating inflammatory cytokines, whereas NLR reflects the distribution of circulating immune-cell populations and may additionally integrate neurohormonal and physiological stress (10,12,14). Their differing associations may therefore reflect distinct biological domains, differences in temporal kinetics, or the limited sample size of the CRP subgroup. Larger studies incorporating high-sensitivity CRP, interleukin-6, tumor necrosis factor pathways, neutrophil activation markers, and other immune phenotypes will be required to determine which inflammatory pathways most closely track pulmonary vascular disease in HFpEF.

### Potential Upstream Hematopoietic Mechanisms

Age-related alterations in hematopoiesis provide one potential, although currently speculative, link between leukocyte phenotype and cardiovascular inflammation. Clonal hematopoiesis of indeterminate potential (CHIP) has been associated with incident and adverse heart failure outcomes and may amplify innate immune signaling (28,29). Individuals with clonal hematopoiesis have also been reported to exhibit higher NLR values (30). These observations raise the hypothesis that age-related hematopoietic reprogramming may contribute to inflammatory heterogeneity within HFpEF. However, genomic profiling was not performed in the present cohort, and no inference regarding clonal hematopoiesis or other specific upstream inflammatory mechanisms can be made from these data. Future studies integrating leukocyte genomics, immune phenotyping, and invasive cardiopulmonary physiology could address this possibility.

### Clinical and Research Implications

NLR has several characteristics that make the present findings potentially clinically relevant: it is inexpensive, routinely available, reproducible from standard complete blood counts, and requires no dedicated biomarker assay. However, the present study does not establish an NLR threshold for clinical decision-making, nor does it support the use of NLR as a substitute for invasive hemodynamic assessment. Rather, NLR may represent a readily accessible signal that identifies patients with HFpEF-PH in whom greater pulmonary vascular burden and adverse cardiovascular risk coexist.

This distinction may become important as PH associated with left heart failure moves toward more granular phenotyping and therapeutic enrichment strategies(9). Trials directed broadly at PH associated with left heart disease have historically been disappointing, potentially in part because hemodynamically and biologically heterogeneous populations were studied together. Contemporary approaches increasingly emphasize identification of patient subsets with specific pulmonary vascular abnormalities and potentially modifiable biological pathways (9). Whether an inflammatory phenotype characterized by elevated NLR identifies a subgroup with distinct treatment responsiveness remains unknown. Prospective studies incorporating serial NLR measurements, comprehensive inflammatory profiling, exercise hemodynamics, RV-pulmonary arterial coupling, and response to HFpEF-directed or anti-inflammatory therapies will be required before NLR can be considered an actionable biomarker.

### Study Limitations

Several limitations should be acknowledged. First, this was a retrospective, single-center study restricted to patients undergoing clinically indicated RHC, introducing referral and selection bias and limiting generalizability to the broader HFpEF population, particularly patients who do not undergo invasive hemodynamic evaluation. The modest sample size also limited extensive multivariable and subgroup analyses.

Second, NLR and invasive hemodynamics were assessed at a single time point; NLR may be influenced by acute illness, medications, renal dysfunction, and other systemic conditions, and residual confounding cannot be excluded despite exclusion of major hematologic disorders. Third, the cross-sectional association between NLR and hemodynamics does not establish causality, and elevated PVR reflects pulmonary vascular load rather than direct evidence of structural vascular remodeling. Fourth, PVR classification may be affected by variability in PAWP and cardiac output measurements, particularly around the 2-Wood-unit threshold. Finally, CRP was available only in a subset of patients, comprehensive inflammatory profiling was unavailable, and longitudinal changes in HFpEF therapies were not systematically captured. These findings therefore require validation in larger prospective cohorts incorporating serial inflammatory assessment and standardized invasive hemodynamic phenotyping.

## Conclusions

In patients with HFpEF and invasively confirmed postcapillary pulmonary hypertension, higher NLR was associated with greater pulmonary vascular hemodynamic burden, a higher likelihood of CpcPH, and increased cardiovascular risk. The association between NLR and pulmonary vascular load occurred without a corresponding significant increase in left-sided filling pressure, suggesting that NLR identifies information not captured by congestion alone. These findings position NLR as a readily available marker of a high-risk HFpEF-PH phenotype rather than a direct measure of pulmonary vascular remodeling. Prospective studies integrating longitudinal inflammatory profiling with comprehensive pulmonary vascular and right ventricular phenotyping are warranted to determine the biological basis, reproducibility, and potential therapeutic relevance of this association.

## Data Availability

The data underlying this study are not publicly available because of patient privacy and institutional restrictions. Deidentified data may be made available from the corresponding author upon reasonable request and subject to institutional and regulatory approval.

## Acknowledgements

ChatGPT (OpenAI) was used for editorial assistance in refining the language, organization, and presentation of the manuscript. All scientific content, statistical analyses, interpretations, references, and final wording were critically reviewed and approved by the authors, who take full responsibility for the manuscript.

## Nonstandard Abbreviations and Acronyms

CpcPH: combined post- and precapillary pulmonary hypertension
HFpEF: heart failure with preserved ejection fraction
IpcPH: isolated postcapillary pulmonary hypertension
NLR: neutrophil-to-lymphocyte ratio
PAWP: pulmonary artery wedge pressure
PVR: pulmonary vascular resistance
RAP: right atrial pressure
RHC: right heart catheterization
TPG: transpulmonary pressure gradient

